# Primary Blastic Plasmacytoid Dendritic Cell Neoplasm: a US Population-Based Study

**DOI:** 10.1101/2023.01.05.23284215

**Authors:** Fan Wang

## Abstract

**Background:** Blastic plasmacytoid dendritic cell neoplasm (BPDCN) is a rare and poorly understood hematopoietic malignancy. This study aimed to investigate the clinical characteristics and prognostic factors in patients with primary BPDCN.

**Methods:** Patients diagnosed with primary BPDCN from 2000 to 2019 were extracted from the Surveillance, Epidemiology and End Results (SEER) database. Independent prognostic factors were evaluated based on the univariate and multivariate Cox regression analysis. The nomogram was then constructed to predict the overall survival of primary BPDCN patients at 3, 5 and 10 years after diagnosis.

**Results:** A total of 668 primary BPDCN patients were included in this study. The average age was 35.7 ± 25.4 years, with 68.7% being male. The mostly affected sites were lymph nodes (59.9%). Most patients (69.9%) received chemotherapy or radiation therapy. For all the patients, the 1-year, 3-year, 5-year, and 10-year overall survival (OS) were 78.0%, 62.6%, 59.0%, and 56.3%, respectively, and the corresponding disease-specific survival (DSS) were 80.6%, 66.8%, 63.5, and 61.8%, respectively. Multivariate cox analysis indicated that age and marital status of other (divorced, widowed and separated) at diagnosis were independent prognostic factors for DSS, but only age was for OS. The 5-year OS rate significantly declined with increasing age: age <15, 89.3%; age 15-39, 57.9%; age 40-64, 51.2%; age ≥65, 26.1%. Nomograms were further constructed to predict the possibility of OS and DSS with good performances.

**Conclusions:** Primary BPDCN is a rare disease, age and marital status were associated with survival of primary BPDCN patients, and age was an independent prognostic factor for OS.

## Introduction

Blastic plasmacytoid dendritic cell neoplasm (BPDCN) is a rare hematopoietic malignancy characterized by aggressive proliferation of precursor plasmacytoid dendritic cells^1^. It is estimated that BPDCN may represent 0.44% of all hematologic malignancies^2^. BPDCN occurs in all races and geographic areas^3^. It affects all age groups, but most patients are adults, especially older adults, with a median age at diagnosis of 65 to 67 years^3^. BPDCN shows a moderate male predominance with an approximate male-to-female ratio of 2.5:1^4^. The most common presentation of BPDCN is asymptomatic skin lesions, other sites including lymph nodes and bone marrow (BM) are also involved often due to systemic dissemination^5^. The diagnosis of BPDCN needs the presence of at least 4 of the following 6 antigens: CD4, CD56, CD303/BDCA-2, TCL-1, CD2AP and CD123, along with the absence of lineage-specific markers^1^. Optimal treatment for BPDCN has not been well defined^6^. Traditional chemotherapy regimens modeled after initial induction therapies for acute leukemia and lymphoma can achieve a complete remission (CR) rate of 70% to 90%^7^. However, the remissions are mostly short-lived, giving rise to a disappointing median overall survival (OS) of approximate 8 to 12 months^8^. Studies indicate that BPDCN patients who received allogeneic hematopoietic stem cell transplantation (allo-HSCT) during first complete remission had better overall survival compared with those who received chemotherapy alone (median OS, 22.7 months vs 7.1 months) ^9^. Up to date, BPDCN still remains poorly understood owing to its rarity. In this study, we aim to delineate the factors affecting the survival of primary BPDCN patients based on a population-based study using the national Cancer Institute’s Surveillance, Epidemiology and End Results (SEER) database.

## Materials and Methods

### Data collection

Data for the current study were collected from the Surveillance, Epidemiology, and End Results (SEER) Program (https://seer.cancer.gov/) maintained by the National Cancer Institute (NCI). The SEER*Stat software version 8.4.0.1 (https://seer.cancer.gov/seerstat/, accessed on November 2, 2022) was used to retrieve the data. Through the “Incidence-SEER Research Plus Data, 17 Registries, Nov 2021 Sub (2000-2019),” patients diagnosed with BPDCN between 2000 and 2019 were selected using the case listing session and cases with only known age (censored at age 89 years) and only malignant behavior were taken in. As shown in the flow chart (Figure 1), the inclusion criteria of BPDCN patients were as follows: (1) the International Classification of Diseases for Oncology (ICD-O-3) histologic code (9727/3); (2) the sequence number indicating the only primary or first primary; (3) the patient’s survival time was 0 or unknown. The exclusion criteria were as follows: (1) the type of reporting source was “death certificate only”; (2) the diagnosis confirmation was unknown. A total of 668 patients with primary BPDCN were included in the final cohort. Since the SEER data used in this study is publicly available and the patient’s private information was anonymous and cannot be reidentified, the ethical approval from the ethics committee was not required.

**Figure 1.**
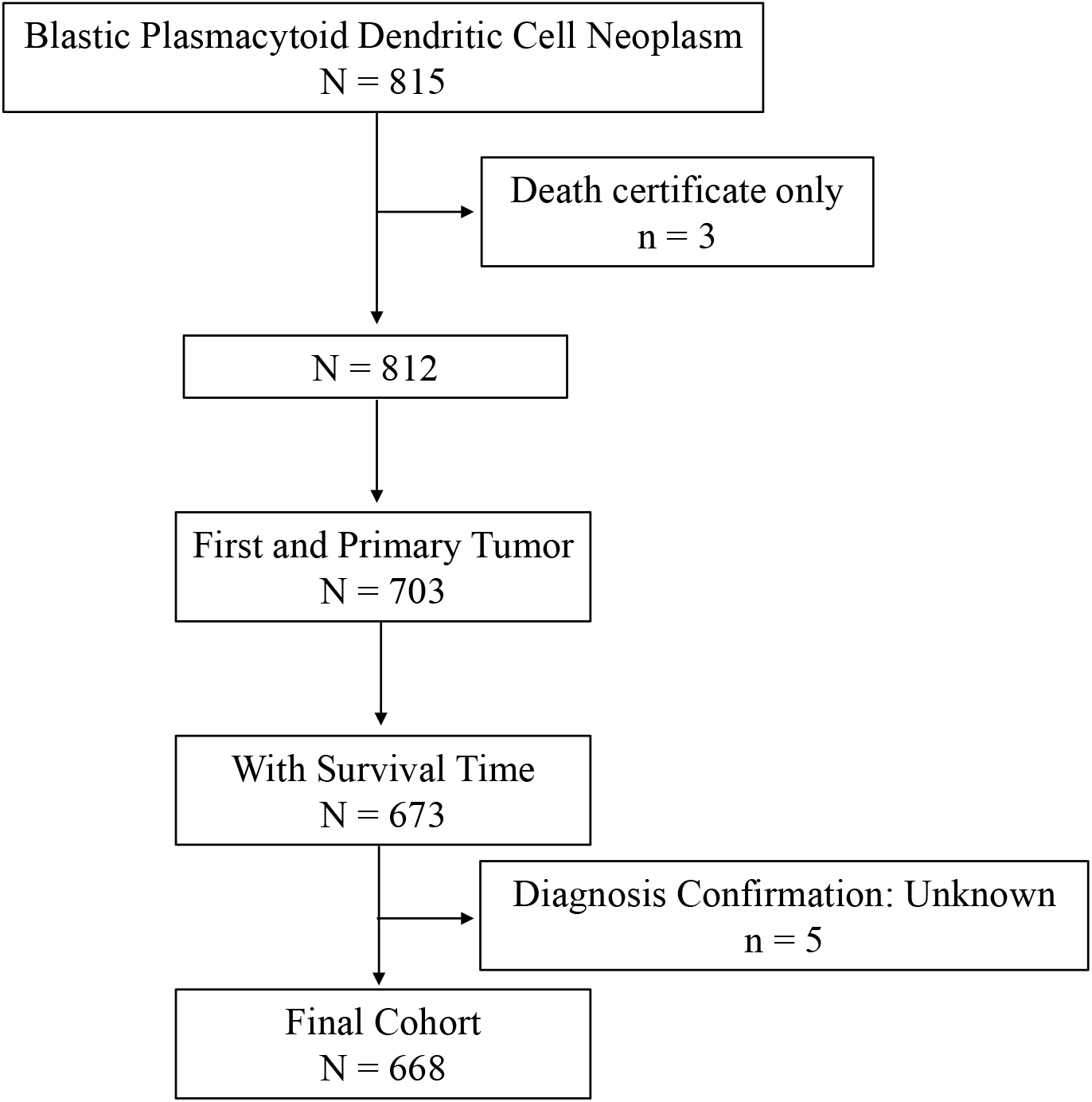
Flow chart of study cohort selection using the SEER database. A flow diagram of BPDCN patient selection in this study. BPDCN, blastic plasmacytoid dendritic cell neoplasm; SEER, Surveillance, Epidemiology, and End Results.

### Variable Definition

The variables collected for analysis included age, sex, race, marital status, year of diagnosis, primary site, vital status, survival months, COD to site recode, cause-specific death classification, cause of death to site, primary site, sequence number, first malignant primary indicator, total number of in situ/malignant tumors for patient, type of reporting source, diagnostic confirmation, chemotherapy recode, and radiation recode. Age was defined as the age attained at diagnosis of BPDCN and were categorized into 4 groups: < 15 years old, 15–39 years old, 40–64 years old, and 60 + years old. Race were classified into African American, White, and Other (Asian/Pacific Islander, American Indian/Alaska Native, Unknown). Marital status was grouped into married, single, and other (including “Divorced”, “Separated”, “Widowed” and “Unknown”). Therapy was categorized into Chemo/RT (chemotherapy or radiation therapy), Chemo + RT (chemotherapy and radiation therapy), No/Unknown. The variable “sequence number” was classified into two groups: one primary only and with SPMs (1st of 2 or more primaries, indicating patients had second primary malignancies other than BPDCN). Cause-of-death information was taken from the “COD to site recode” field. BPDCN-related death was defined in the SEER database as “dead (attributed to this cancer diagnosis)”. The definition of BPDCN-unrelated death was defined in the SEER database as death “dead (attributable to causes other than this cancer diagnosis)”. Overall survival time was calculated from the date of BPDCN diagnosis to death or last follow up.

### Statistical analysis

All statistical analysis of the present study was performed using the R program language (http://www.r-project.org/, version 4.2.1; R Foundation for Statistical Computing, Vienna, Austria). Baseline characteristics of primary BPDCN patients were compared between two groups: “alive or BPDCN-unrelated death” and “BPDCN-related death”. Comparisons of continuous data and categorical data were performed using Student’s *t*-test and chi-square test, respectively. The overall survival (OS) and disease-specific survival (DSS) were estimated with the Kaplan-Meier method using the log-rank tests. To determine the independent prognostic factors of primary BPDCN patients, univariate and multivariate Cox proportional regression analyses were performed, the hazards ratio (HR) estimates and 95% confidence intervals (95% CIs) were reported. Preliminary evaluation of Cox proportional hazards regression by proportional-hazards assumption test and Schoenfeld residual test revealed not violation of proportion assumption in all variables. Variables with *P*□<□0.05 in the univariate Cox proportional hazard model were further analyzed in the multivariate Cox proportional hazard model. Based on independent prognostic factors from the multivariate Cox analysis, nomograms were constructed to predict the 3-, 5- and 10-year OS and DSS probabilities. The time-dependent receiver operating characteristic (ROC) curves of nomograms at 3-, 5- and 10-year were generated, and the corresponding area under the curve (AUC) were applied to assess the discrimination. In addition, the calibration curves and decision curve analysis (DCA) nomograms at 3-, 5- and 10-year were plotted to evaluate the performance of the nomogram. All *P* values were two-sided and a *P* value < 0.05 was defined as statistically significant.

## Results

### Baseline Characteristics of Primary BPDCN Patients

As depicted in Figure 1, a total 668 patients were finally identified as primary BPDCN in the SEER 17 registry, Nov 2021 Sub (2000-2019) from January 2020 to December 2019. The primary site distribution analysis showed that the mostly affected sites were lymph nodes (*n*=400, 59.9%), bone marrow (*n*=111, 16.6%), skin (*n*=74, 11.1%) and mediastinum (*n*=23, 3.4%). The other sites such as nervous system, testis, lung, spleen, bones, liver, kidney, heart, and colon were rarely affected. A detailed description of primary site distribution of BPDCN was shown in Table 1. For all patients in the study cohort, 68.7% were male, more than twice that of female (*n*=209, 31.3%; Table 2). The average age at diagnosis was 35.7 ± 25.4 years, age distribution was as follows: < 15 years old (28.0%), 15-39 years old (30.7%), 40-64 years old (21.0%), and 60+ years old (20.4%). The majority of primary BPDCN patients were white (81.6%), African American and Other (including Asian/Pacific Islander, American Indian/Alaska Native and Unknown) occupied 10.3% and 8.1%, respectively. Most of the patients were single who had never been married (52.8%) at diagnosis, followed by married (36.4%) and other marital status (10.8%) including “Divorced”, “Separated”, “Widowed” and “Unknown”. The highest incidence of year range group was 2000-2004 (41.6%). For all primary BPDCN cases in this study, 8.5% had second primary malignancies (SPMs). In general, 69.9% primary BPDCN patients received chemotherapy or radiation therapy, followed by 19.6% patients treated with chemotherapy combined with radiation therapy. At the time of last follow-up, 385 (57.6%) patients were alive; 233 (34.9%) deaths were attributable to primary BPDCN, and an additional 50 (7.5%) patients died due to other causes such as diseases of heart (n=13, 26.0%), pulmonary disease (n=2, 4.0%), septicemia (n=2, 4.0%), cerebrovascular diseases (n=1, 2.0%), kidney diseases (n=2, 4.0%), melanoma of the skin (n=1, 2.0%) and suicide and self-inflicted injury (n=3, 6.0%). Age (*P* < 0.001) and marital status (*P* < 0.001) were found to be significantly different between the two survival groups “alive or unrelated death” and “BPDCN-related death”. The age group “65+ “ had the most cases of “BPDCN-related death”(n=79, 33.9%). The epidemiologic characteristics and survival comparison were summarized in Table 2.

**Table 1.** Distribution of the primary sites of blastic plasmacytoid dendritic cell neoplasm

| Primary Sites | Overall<br>(N=668) |
| --- | --- |
| Accessory sinus, NOS | 1 (0.1%) |
| Bladder, NOS | 1 (0.1%) |
| **Bone marrow** | 111 (16.6%) |
| Central portion of breast | 1 (0.1%) |
| Cervix uteri | 1 (0.1%) |
| Colon, NOS | 1 (0.1%) |
| Connective, subcutaneous | 13 (1.9%) |
| Fundus of stomach | 1 (0.1%) |
| Head of pancreas | 1 (0.1%) |
| Heart | 1 (0.1%) |
| Jejunum | 1 (0.1%) |
| Kidney, NOS | 2 (0.3%) |
| Lateral wall of bladder | 1 (0.1%) |
| Liver | 1 (0.1%) |
| Long bones | 4 (0.6%) |
| **Lymph nodes** | 400 (59.9%) |
| Lymph node, NOS | 75 (11.2%) |
| Lymph nodes of axilla or arm | 3 (0.4%) |
| Lymph nodes of head, face & neck | 32 (4.8%) |
| Lymph nodes of inguinal region or leg | 6 (0.9%) |
| Abdominal lymph nodes | 9 (1.3%) |
| Intrathoracic lymph nodes | 77 (11.5%) |
| Pelvic lymph nodes | 4 (0.6%) |
| Waldeyers ring | 1 (0.1%) |
| Lymph nodes of multiple regions | 193 (28.9%) |
| Maxillary sinus | 3 (0.4%) |
| **Mediastinum** | 23 (3.4%) |
| Anterior mediastinum | 13 (1.9%) |
| Mediastinum, NOS | 10 (1.5%) |
| **Nervous system** | 4 (0.6%) |
| Brain, NOS | 2 (0.3%) |
| Spinal cord | 1 (0.1%) |
| Nervous system, NOS | 1 (0.1%) |
| Orbit, NOS | 2 (0.3%) |
| Ovary | 1 (0.1%) |
| **Overlapping lesion** | 4 (0.6%) |
| Overlapping lesion of brain | 1 (0.1%) |
| Overlapping lesion of brain & CNS | 1 (0.1%) |
| Overlapping lesion of breast | 1 (0.1%) |
| Overlapping lesion of skin | 1 (0.1%) |
| Parotid gland | 1 (0.1%) |
| Pelvic bones, sacrum, coccyx and associated joints | 1 (0.1%) |
| Pharynx, NOS | 1 (0.1%) |
| **Skin** | 74 (11.1%) |
| Skin of lower limb and hip | 10 (1.5%) |
| Skin of scalp and neck | 4 (0.6%) |
| Skin of trunk | 18 (2.7%) |
| Skin of upper limb and shoulder | 9 (1.3%) |
| Skin other/unspec parts of face | 9 (1.3%) |
| Skin, NOS | 24 (3.6%) |
| Small intestine, NOS | 2 (0.3%) |
| Spleen | 1 (0.1%) |
| Testis | 3 (0.4%) |
| Tonsil | 2 (0.3%) |
| Unknown primary site | 1 (0.1%) |
| Upper lobe, lung | 2 (0.3%) |
| Vertebral column | 2 (0.3%) |

Abbreviations: NOS, not otherwise specified.

**Table 2.** Comparison of patient baseline characteristics between BPDCN-related death and alive or unrelated death in the primary BPDCN cohort.

| Characteristics | Total<br>(N=668) | Alive or<br>Unrelated<br>Death<sup>5</sup><br>(N=435) | BPDCN-Related<br>Death<sup>6</sup><br>(N=233) | P-value |
| --- | --- | --- | --- | --- |
| Sex, n (%) |  |  |  |  |
| Male | 459 (68.7%) | 292 (67.1%) | 167 (71.7%) | 0.482 |
| Female | 209 (31.3%) | 143 (32.9%) | 66 (28.3%) |  |
| Age, n (%) |  |  |  |  |
| <15 | 187 (28.0%) | 170 (39.1%) | 17 (7.3%) | <0.001 |
| 15-39 | 205 (30.7%) | 129 (29.7%) | 76 (32.6%) |  |
| 40-64 | 140 (21.0%) | 79 (18.2%) | 61 (26.2%) |  |
| 65+ | 136 (20.4%) | 57 (13.1%) | 79 (33.9%) |  |
| Race, n (%) |  |  |  |  |
| White | 545 (81.6%) | 355 (81.6%) | 190 (81.5%) | 0.702 |
| African American | 69 (10.3%) | 41 (9.4%) | 28 (12.0%) |  |
| Other<sup>1</sup> | 54 (8.1%) | 39 (9.0%) | 15 (6.4%) |  |
| Marital Status, n (%) |  |  |  |  |
| Married | 243 (36.4%) | 127 (29.2%) | 116 (49.8%) | <0.001 |
| Single<sup>2</sup> | 353 (52.8%) | 278 (63.9%) | 75 (32.2%) |  |
| Other<sup>3</sup> | 72 (10.8%) | 30 (6.9%) | 42 (18.0%) |  |
| Diagnosis Year, n (%) |  |  |  |  |
| 2000-2004 | 278 (41.6%) | 167 (38.4%) | 111 (47.6%) | 0.0952 |
| 2005-2009 | 130 (19.5%) | 94 (21.6%) | 36 (15.5%) |  |
| 2010-2014 | 118 (17.7%) | 71 (16.3%) | 47 (20.2%) |  |
| 2015-2019 | 142 (21.3%) | 103 (23.7%) | 39 (16.7%) |  |
| **Sequence Number, *n* (%)** |  |  |  |  |
| One primary only | 611 (91.5%) | 396 (91.0%) | 215 (92.3%) | 0.861 |
| With SPMs<sup>4</sup> | 57 (8.5%) | 39 (9.0%) | 18 (7.7%) |  |
| **Therapy, *n* (%)** |  |  |  |  |
| Chemo/RT | 467 (69.9%) | 310 (71.3%) | 157 (67.4%) | 0.401 |
| Chemo + RT | 131 (19.6%) | 87 (20.0%) | 44 (18.9%) |  |
| No/Unknown | 70 (10.5%) | 38 (8.7%) | 32 (13.7%) |  |

<sup>1</sup> Races including Asian/Pacific Islander, American Indian/Alaska Native and Unknown.
<sup>2</sup> Marital status at diagnosis was single (never married).
<sup>3</sup> Marital status of divorced, widowed and separated at diagnosis.
<sup>4</sup> Sequence number of “1st of 2 or more primaries”, indicating BPDCN patients had second primary malignancies other than BPDCN.
<sup>5</sup> Alive or Dead of other cause such as diseases of heart, cerebrovascular diseases, septicemia, and so on.
<sup>6</sup> Death attributable to BPDCN.
Abbreviations: BPDCN, blastic plasmacytoid dendritic cell neoplasm; Chemo, chemotherapy; COD, cause of death; RT, radiation therapy; SPMs, second primary malignancies.

### Survival Analysis of Primary BPDCN Patients

There were 283 deaths during the follow-up period, and 233 deaths were disease specific. As shown in Figure 2A and B, the 1-year, 3-year, 5-year, and 10-year OS were 78.0%, 62.6%, 59.0%, and 56.3%, respectively, and the corresponding DSS were 80.6%, 66.8%, 63.5, and 61.8%, respectively. Although there seemed to be significant differences for OS between the diagnosis years (*P* = 0.027; Figure 2C), but pairwise analysis with *P* value adjustment using the “Benjamini-Hochberg” method showed that they were not significantly different (Figure 2C). There were neither significant difference for DSS between the diagnosis years (*P* = 0.064; Figure 2D).

**Figure 2.**
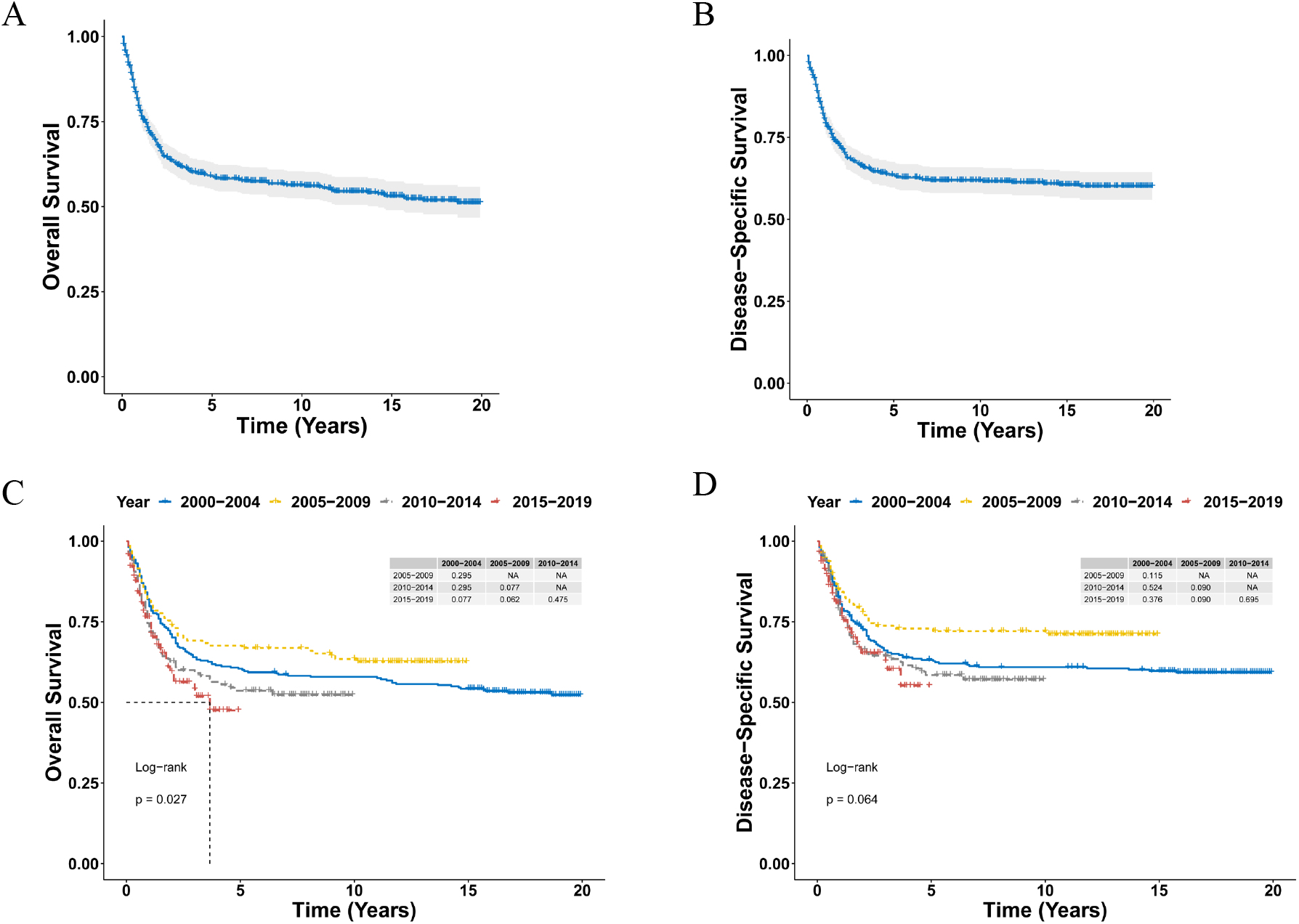
Survival analysis of primary BPDCN. **(A, B)** OS (A) and DSS (B) curves for all primary BPDCN patients. **(C, D)** Survival curves of OS (E) and DSS (F) according to the diagnosis years. BPDCN, blastic plasmacytoid dendritic cell neoplasm; OS, overall survival; DSS, disease-specific survival.

Furthermore, Kaplan-Meier analysis of OS suggested older age (*P* < 0.0001; Figure 3A), male (*P* = 0.049; Figure 3B), unmarried patients (*P* < 0.0001; Figure 3D), and without treatments (*P* = 0.0007; Figure 3F) were associated with poorer overall survival. The 5-year OS rate significantly declined with increasing age: <15, 89.3%; 15-39, 57.9%; 40-64, 51.2%; ≥65, 26.1% (*P* < 0.0001; Figure 3A). There was no significant difference for overall survival between the race categories (*P* = 0.53, Figure 3C), and the overall survival was also not significantly different between the BPDCN only group and the group with SPMs (*P* = 0.67, Figure 3E). Moreover, as shown in Figure 4, Kaplan-Meier analysis of DSS showed similar results, except that there was no significant difference of DSS between male and female (*P* = 0.17, Figure 4B).

**Figure 3.**
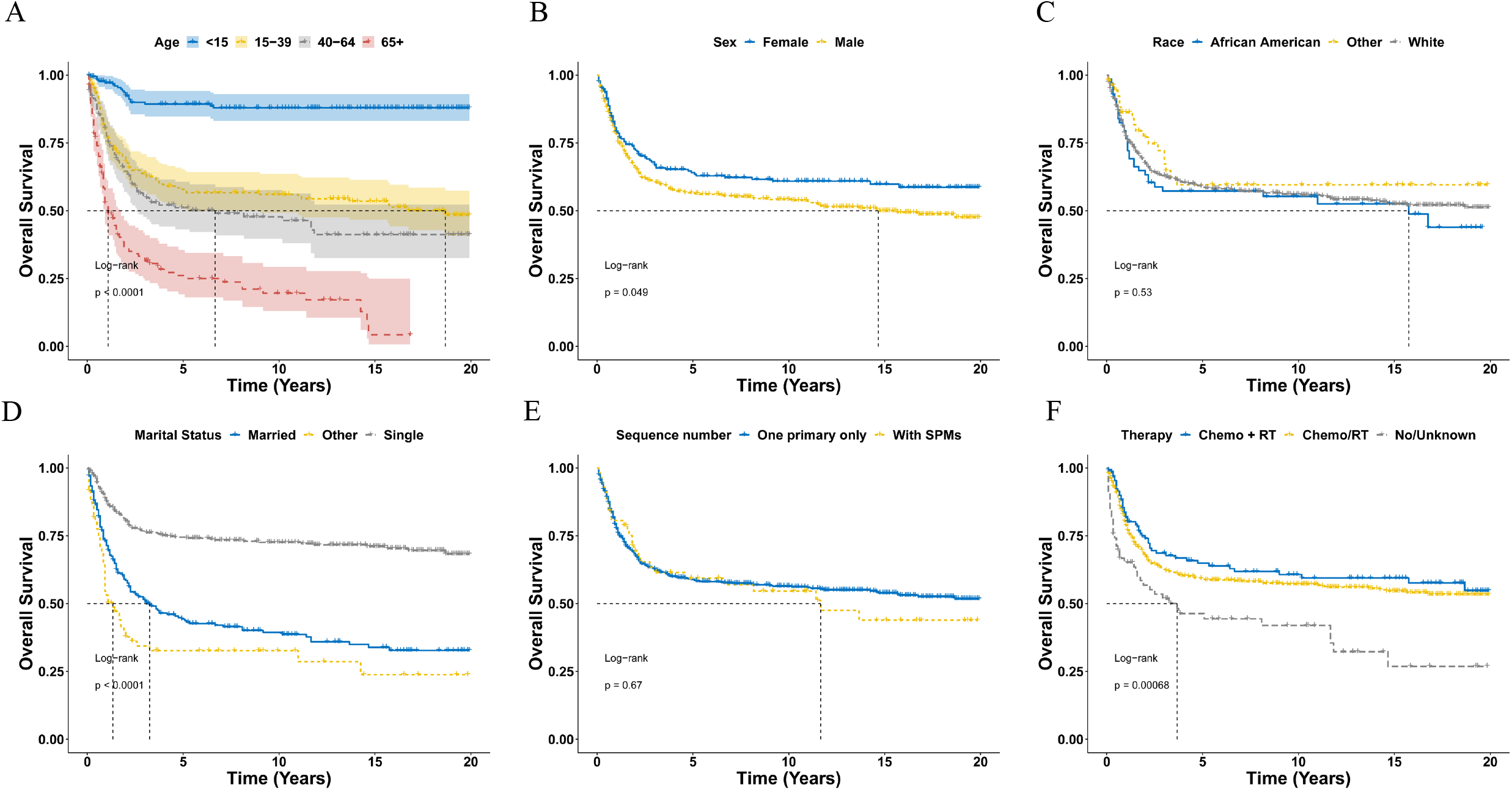
Overall survival analysis of primary BPDCN stratified by age (A), sex (B), race (C), and marital status (D), sequence number (E) and therapy (F) using Kaplan-Meier method. BPDCN, blastic plasmacytoid dendritic cell neoplasm; Chemo, chemotherapy; RT, radiation therapy; SPMs, second primary malignancies.

**Figure 4.**
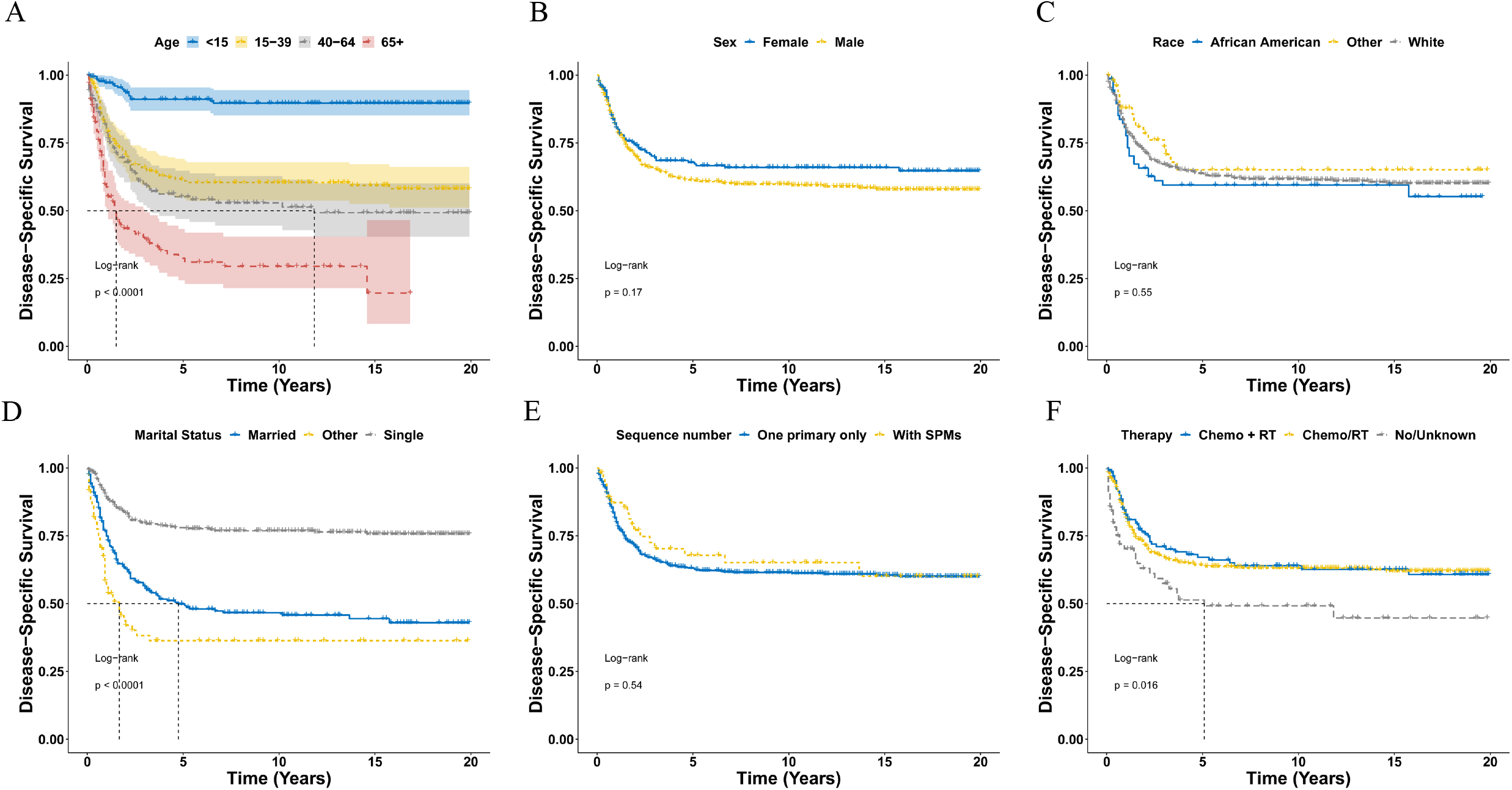
Disease-specific survival analysis of primary BPDCN stratified by age (A), sex (B), race (C), and marital status (D), sequence number (E) and therapy (F) using Kaplan-Meier method. BPDCN, blastic plasmacytoid dendritic cell neoplasm; Chemo, chemotherapy; RT, radiation therapy; SPMs, second primary malignancies.

### Univariate and Multivariable Cox Regression Analysis of Primary BPDCN Patients

The univariate cox regression analysis of OS showed that older age, male, marital status of single and other (divorced, widowed and separated) at diagnosis, without therapy were significantly associated with worse OS (Table 3), while the other variables such as sex, race and sequence number did not affect the overall survival outcomes of primary BPDCN. Similar results were obtained with the univariate cox regression analysis of DSS (Table 3). These significant variables were further included for the multivariate cox analysis. Multivariace Cox analysis indicated that only age was an independent prognostic factor for OS (Table 4). Regarding DSS, age and marital status of other (divorced, widowed and separated) at diagnosis were identified as independent prognostic factors (Table 4).

**Table 3.** Univariable cox regression analysis for OS and DSS of primary BPDCN patients

| Parameters | OS |  |  | DSS |  |  |
| --- | --- | --- | --- | --- | --- | --- |
|  | HR | 95% CI | P-value | HR | 95% CI | P-value |
| Sex |  |  |  |  |  |  |
| Male | ref |  |  | ref |  |  |
| Female | 0.77 | [0.59, 1.00] | 0.05 | 0.82 | [0.62, 1.09] | 0.18 |
| Age |  |  |  |  |  |  |
| <15 | ref |  |  | ref |  |  |
| 15-39 | 5.18 | [1.01, 1.03] | < 0.001 | 4.99 | [2.95, 8.44] | < 0.001 |
| 40-64 | 6.54 | [3.98, 10.77] | < 0.001 | 6.27 | [3.66, 10.75] | < 0.001 |
| 65+ | 14.54 | [8.95, 23.61] | < 0.001 | 12.29 | [7.25, 20.83] | < 0.001 |
| Race |  |  |  |  |  |  |
| White | ref |  |  | ref |  |  |
| African American | 1.11 | [0.77, 1.60] | 0.58 | 1.15 | [0.77, 1.70] | 0.50 |
| Other<sup>1</sup> | 0.80 | [0.49, 1.29] | 0.35 | 0.81 | [0.48, 1.36] | 0.42 |
| Marital Status |  |  |  |  |  |  |
| Married | ref |  |  | ref |  |  |
| Single<sup>2</sup> | 0.35 | [0.27, 0.46] | < 0.001 | 0.35 | [0.26, 0.47] | < 0.001 |
| Other<sup>3</sup> | 1.46 | [1.05, 2.02] | 0.02 | 1.53 | [1.08, 2.18] | 0.02 |
| Diagnosis Year |  |  |  |  |  |  |
| 2000-2004 | ref |  |  | ref |  |  |
| 2005-2009 | 0.82 | [0.59, 1.15] | 0.25 | 0.69 | [0.49, 0.97] | 0.03 |
| 2010-2014 | 1.24 | [0.90, 1.71] | 0.19 | 0.61 | [0.61, 1.18] | 0.32 |
| 2015-2019 | 1.44 | [1.03, 2.02] | 0.04 | 0.79 | [0.56, 1.13] | 0.20 |
| Sequence Number |  |  |  |  |  |  |
|  | HR | 95% CI | P-value | HR | 95% CI | P-value |
| One primary only | *ref* |  |  | *ref* |  |  |
| With SPMs<sup>4</sup> | 1.09 | [0.73, 1.62] | 0.67 | 0.86 | [0.53, 1.39] | 0.53 |
| **Therapy** |  |  |  |  |  |  |
| Chemo/RT | *ref* |  |  | *ref* |  |  |
| Chemo + RT | 0.87 | [0.63, 1.18] | 0.37 | 0.96 | [0.69, 1.34] | 0.82 |
| No/Unknown | 1.80 | [1.29, 2.53] | < 0.001 | 1.70 | [1.16, 2.49] | 0.01 |

<sup>1</sup>Races including Asian/Pacific Islander, American Indian/Alaska Native and Unknown.
<sup>2</sup> Marital status at diagnosis was single (never married).
<sup>3</sup> Marital status of divorced, widowed and separated at diagnosis.
<sup>4</sup> Sequence number of “1st of 2 or more primaries”, indicating BPDCN patients had second primary malignancies other than BPDCN.
Abbreviations: BPDCN, blastic plasmacytoid dendritic cell neoplasm; Chemo, chemotherapy; CI, confidence interval; DSS, disease-specific survival; HR, hazard ration; OS, overall survival; RT, radiation therapy; SPMs, second primary malignancies.

**Table 4.** Multivariable cox regression analysis for OS and DSS of primary BPDCN patients

| Parameters | OS |  |  | DSS |  |  |
| --- | --- | --- | --- | --- | --- | --- |
|  | HR | 95% CI | P-value | HR | 95% CI | P-value |
| **Age** |  |  |  |  |  |  |
| <15 | *ref* |  |  | *ref* |  |  |
| 15-39 | 4.95 | [3.00, 8.20] | < 0.001 | 4.52 | [2.61, 7.80] | < 0.001 |
| 40-64 | 5.89 | [3.35, 10.40] | < 0.001 | 5.17 | [2.79, 9.60] | < 0.001 |
| 65+ | 12.58 | [7.11, 22.20] | < 0.001 | 9.77 | [5.21, 18.30] | < 0.001 |
| **Marital Status** |  |  |  |  |  |  |
| Married | *ref* |  |  | *ref* |  |  |
| Single<sup>1</sup> | 0.92 | [0.66, 1.30] | 0.62 | 0.84 | [0.58, 1.20] | 0.35 |
| Other<sup>2</sup> | 1.39 | [1.00, 1.90] | 0.05 | 1.47 | [1.03, 2.10] | 0.03 |
| **Therapy** |  |  |  |  |  |  |
| Chemo/RT | *ref* |  |  | *ref* |  |  |
| Chemo + RT | 0.91 | [0.66, 1.30] | 0.57 | 1.00 | [0.71, 1.40] | 0.99 |
| No/Unknown | 0.94 | [0.66, 1.30] | 0.74 | 0.91 | [0.61, 1.30] | 0.64 |

<sup>1</sup> Marital status at diagnosis was single (never married).
<sup>2</sup> Marital status of divorced, widowed and separated at diagnosis.
Abbreviations: BPDCN, blastic plasmacytoid dendritic cell neoplasm; Chemo, chemotherapy; CI, confidence interval; DSS, disease-specific survival; HR, hazard ration; OS, overall survival; RT, radiation therapy.

### Establishment and Evaluation of Nomogram Models

A nomogram was further constructed to predict the primary BPDCN patients’ 3-, 5-, and 10-year OS probability by integrating age and marital status (Figure 5A). The calibration curve of the nomogram showed that the predicted and observed OS probability matched well at 3-, 5-, and 10-year time intervals (Figure 5B). Moreover, the time-dependent ROC curve analyses showed that the 3-year, 5-year and 10-year AUC of the nomogram was 0.738, 0.752 and 0.766, respectively (Figure 5C), indicating the high accuracy of the nomogram models. Additionally, the DCA curves also suggested that the nomograms had good performances to serve as an effective tool for predicting the OS probability (Figure 5D-F). Furthermore, similar results were obtained for the nomograms to predict the 3-, 5-, and 10-year DSS probability of primary BPDCN patients (Supplementary Figure S1 and S2).

**Figure 5.**
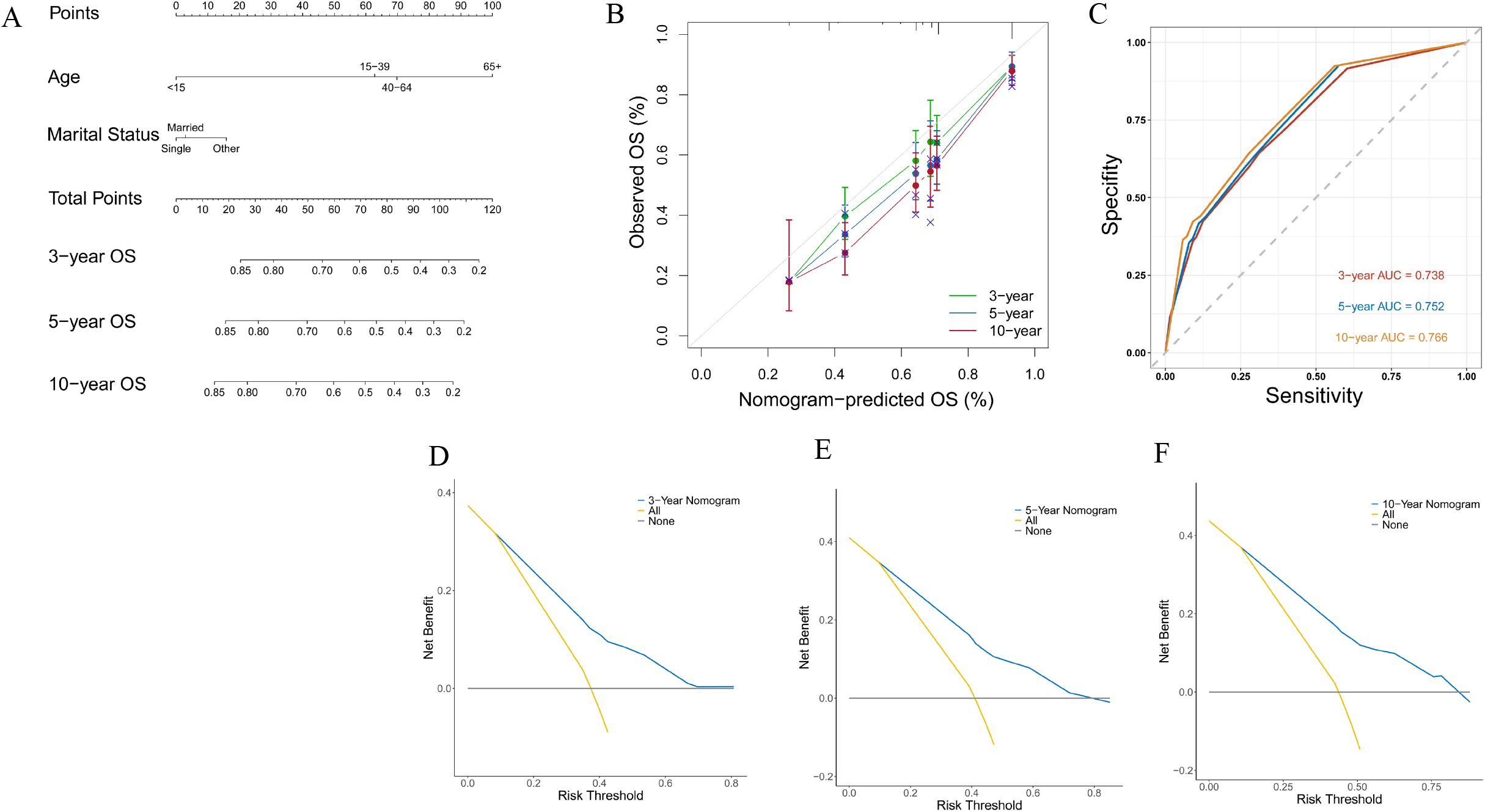
Construction and evaluation of the nomogram of primary BPDCN patients. (A) The nomogram for predicting the probability of 3-, 5-, and 10-year OS. The total points were calculated by integrating scores related to race and marital status and projected to the bottom scale to predict the OS survival probability at 3, 5, and 10 years. (B) The calibration curve of the nomogram for the observed overall survival (OS) probability and predicted OS at 3-year, 5-year, and 10-year. (C) Time-dependent ROC curve analyses of the nomograms for the 3 years, 5 years, and 10 years in the primary BPDCN cohort. (D-F) The decision curve analyses of the nomogram for the 3 years (D), 5 years (E), and 10 years (F) in the primary BPDCN cohort. BPDCN, blastic plasmacytoid dendritic cell neoplasm; OS, overall survival.

## Discussion

The current study identified 668 primary BPDCN patients of 2000-2019 from the SEER database, representing the largest cohort describing the clinical characteristics and outcome of patients with primary BPDCN. The average age was 35.7 ± 25.4 years, predominantly affected male (male: female = 2.2), and the mostly affected sites were lymph nodes (59.9%). Although BPDCN can occur in any age, studies indicated that it was most common in the elderly patients, with a median age of 61-67 years^10^, which differed from our results. The variable demographic characteristics of primary BPDCN in current study may reflect the different age distribution and the rarity and heterogeneity of BPDCN.

Previous studies have demonstrated that age was significantly correlated with the prognosis of BPDCN^1,3,4^. In the current study, worse OS and DSS were significantly associated with older age, the 5-year OS and DSS rate remarkably declined with increasing age, especially for patients ≥65 years of age, which was consistent with prior studies indicating that pediatric BPDCN patients had more favorable clinical outcomes^8^. Further univariate and multivariate cox analysis confirmed that age was an independent negative prognostic factor for OS and DSS for primary BPDCN patients, which may be partially associated with the decreased intensity of chemotherapies and inability for allo-HSCT as a result of aging^1,4,7^.

Marital status has been increasingly recognized as an important prognostic factor for cancer patients^11-13^. Our study suggested a 1.47-fold hazard ratio in divorced/widowed/separated primary BPDCN patients compared to married and single ones after multivariate cox analysis by adjusting for other confounding variables. The underlying mechanism of shortened survival in divorced/widowed/separated patients is unclear, may be related with socioeconomic and psychological status since those patients may have experienced more strong fluctuations in socioeconomic and emotional changes.

Up to now, there is no consensus on the optimal therapeutic regime for BPDCN^6^. It has been suggested that traditional leukemia/lymphoma-based chemotherapy regimens did not prolong the overall survival, resulting in only temporary remission, but allo-HSCT especially that performed during the first CR demonstrated durable remissions, with OS rates reaching 74% to 82% at 3 to 4 years^6,7,14-16^. Our study showed that any chemotherapy or radiation therapy was associated with longer OS and DSS compared with no therapy group. However, univariate, and multivariate cox analysis showed that those therapies (including chemotherapy and radiotherapy) had no impact on the survival outcomes of primary BPDCN at all. Although there were numerous introductions of novel drugs such as Tagraxofusp and Venetoclax for the treatment of BPDCN, which have shown promising results^17,18^. In the current study, based on population analysis of primary BPDCN patients, we found that patients diagnosed between 2015 and 2019 had no significant different OS and DSS than those diagnosed between 2000 and 2004, reflecting the development status of the novel therapies.

The nomogram has become widely used in clinical practice for predicting the prognosis of cancers^19^. In this study, it was found that age and marital status were independent prognostic factors for the survival outcomes of primary BPDCN patients, and the nomograms were constructed to predict 3-, 5-, and 10-year survival based on age and marital status of patients. The calibration curve of the nomogram exhibited good consistency between the predicted and the actual survival probability, and the significantly higher AUC of the nomogram indicating the high accuracy of the nomogram models. Moreover, the DCA curves also suggested that the nomograms had good performances to serve as an effective tool for predicting the OS probability.

There are several limitations with this study. Firstly, other potential prognostic factors such as LDH level, carcinogens exposure, family history, alcohol/smoking consumption history, Epstein-Barr virus status were not documented in the SEER registry, which may have profound effect on the outcome of BPDCN patients.

Secondly, detailed chemotherapy and radiotherapy regimens were not noted in the SEER database, thus it is impossible for us to the analyze the impact of different treatment regimen on the prognosis of BPDCN patients. Thirdly, this is a retrospective study with unavoidable potential biases such as selection bias, recall bias or misclassification bias. Finally, the nomograms of primary BPDCN were constructed and verified by using the same database, which was not further verified by using another independent dataset. Thus, the results of the current study should be interpreted with caution although our study still provided important insights on BPDCN due to the rarity and lacking large-scale trials of the disease.

## Conclusions

In conclusion, primary BPDCN is a rare disease, age, chemotherapy (and/or radiotherapy) and marital status were associated with the survival outcomes of primary BPDCN patients. Older age and marital status of divorced/widowed/separated) at diagnosis were independent worse prognostic factors for DSS, but only age was an independent prognostic factor for OS. A predictive nomogram was also developed to predict the long-term OS of primary BPDCN patients with favorable accuracies. To our knowledge, this is the largest population-based cohort investigating the clinical characteristics and survival outcome of patients with primary BPDCN.

## Data Availability

The data analyzed in this study are from the SEER database (https://seer.cancer.gov/) that are available to the public.

## Acknowledgments

The interpretation of the data is the sole responsibility of the author(s). The author(s) acknowledge the efforts of the National Cancer Institute and the Surveillance, Epidemiology, and End Results (SEER) Program tumor registries in the creation of the SEER database.

## Abbreviations

BPDCN: Blastic Plasmacytoid Dendritic Cell Neoplasm
Chemo: chemotherapy
CI: confidence interval
COD: cause of death
DSS: disease-specific survival
HR: hazard ration
OS: overall survival
RT: radiation therapy
SEER: Surveillance, Epidemiology, and End Results
SPMs: second primary malignancies
NOS: not otherwise specified

## Supplementary Figure Legends

**Figure S1. Construction and evaluation of the nomogram of primary BPDCN patients**. The nomogram for predicting the 3-, 5-, and 10-year DSS probability. The DSS survival rates at 3, 5, and 10 years can be predicted by integrating scores related to race and marital status.

**Figure S2**. (A) The calibration diagram of the nomogram for the observed DSS probability and predicted DSS at 3-year, 5-year, and 10-year. (B) Time-dependent ROC curve analyses of the nomograms in the 3 years, 5 years, and 10 years in the primary BPDCN cohort. (C-E) The decision curve analyses of the nomogram for the 3 years (C), 5 years (D), and 10 years (E) in the primary BPDCN cohort. BPDCN, blastic plasmacytoid dendritic cell neoplasm; DSS, disease-specific survival.

